# Monoallelic *CRMP1* gene variants cause neurodevelopmental disorder

**DOI:** 10.1101/2022.07.05.22276556

**Authors:** Ethiraj Ravindran, Nobuto Arashiki, Lena-Luise Becker, Kohtaro Takizawa, Jonathan Lévy, Thomas Rambaud, Konstantin L. Makridis, Yoshio Goshima, Na Li, Maaike Vreeburg, Bénédicte Demeer, Achim Dickmanns, Alexander P.A Stegmann, Hao Hu, Fumio Nakamura, Angela M. Kaindl

## Abstract

Collapsing response mediator proteins (CRMPs) are key for brain development and function. Here, we link CRMP1 to a neurodevelopmental disorder. We report heterozygous *de novo* variants in the *CRMP1* gene in three unrelated individuals with muscular hypotonia, intellectual disability and/or autism spectrum disorder. Based on *in silico* analysis these variants are predicted to affect the CRMP1 structure. We further analyzed the effect of the variants on the protein structure/levels and cellular processes. We showed that the human *CRMP1* variants are dominant-negative and impact the oligomerization of CRMP1 proteins. Moreover, overexpression of mutant-*CRMP1* variants affect neurite outgrowth of murine cortical neurons. While altered CRMP1 levels have been reported in psychiatric diseases, genetic mutation in *CRMP1* gene has never been linked to human disease. We report for the first-time mutations in the *CRMP1* gene and emphasize its key role in brain development and function by linking directly to a human neurodevelopmental disease.

## Introduction

Neurodevelopment is a fine-tuned process orchestrated by distinct expression and function of several genes and any disturbances in this timely-controlled process culminate in neurodevelopmental disorder.^1,2^ Collapsin response mediator proteins (CRMP) are cytosolic phosphoproteins that are highly and differentially expressed in the nervous system.^3,4^ The five CRMP subtypes (CRMP1-5) form homo- or hetero-tetramers in various combinations and thereby enable distinct functions key for neurodevelopment.^5^ Targeted neurodevelopmental processes include cell migration, axonal outgrowth, dendritic branching, apoptosis mediated through extracellular signaling molecules (Sema3A, reelin, neurotrophins).^6-10^ CRMP function is regulated in a spatiotemporal manner through protein phosphorylation mediated by various kinases such as Cdk5, Rho/ROCK, and GSK3.^11,12^

Given their key function in developmental processes, disturbances in CRMP function can result in neurodevelopmental diseases (NDD). In this line, monoallelic *CRMP5* variants can cause Ritscher-Schinzel syndrome 4 (MIM#619435), a neurodevelopmental disease with craniofacial features, cerebral and cardiovascular malformations, and cognitive dysfunction.^13^ *CRMP4* variants have been associated with amyotrophic lateral sclerosis in the French population.^14^

Here, we link for the first time *CRMP1* variants in three unrelated pedigrees to neurodevelopmental disorder in humans with muscular hypotonia, autism spectrum disorder (ASD) and/or intellectual disability. In humans, maternal CRMP1 autoantibodies have been associated with autism in their children, and increased *CRMP1* mRNA levels were identified in individuals with schizophrenia, attention deficit hyperactivity disorder and ASD.^15,16^ Knockout of *Crmp1* in mice results in schizophrenia-associated behavior, impaired learning and memory, and prepulse inhibition.^17^ On a cellular level, abnormal neurite outgrowth, dendritic development and orientation, and spine maturation of cortical and/or hippocampalneurons have been shown.^7-9^ Loss of *Crmp* affects long-term potentiation maintenance.^9^ In addition, cerebellar development and its loss leads to reduced granule cell proliferation, apoptosis, and migration in the cerebellum of *Crmp1*^-/-^ mice.^10^ Although evidence on altered levels of CRMP1 in neuropsychiatric diseases exists, the mutations in the *CRMP1* gene has not been linked to a human disease.

## Subjects and methods

Written informed consent was obtained from all parents of the patients to participate in this study and publish the data of this research work. The human study adhered to the World Health Association Declaration of Helsinki (2013) and was approved by the local ethics committees of the Charité (approval no. EA1/212/08). All animal experimental protocols were checked and approved by the Institutional Animal Care and Use Committee of the Tokyo Women’s medical University with protocol No. ‘AE21-086’. All animal experiments were performed at daytime. The study was not pre-registered.

### Genetic analyses

In family 1, DNA samples of family members were isolated from peripheral blood lymphocytes, and whole exome sequencing (WES) was performed on a HiSeq XTen Deep Sequencer (Illumina, CA, USA), with an average coverage of ∼36X, according to the manufacturer’s instructions. The primary data analysis was done by a combination of Burrows-Wheeler Alignment (BWA) sequence aligner for reads alignment to human reference genome GRCh37/hg19, Genome Analysis Toolkit (GATK) pipeline for calling single-nucleotide variant (SNV), and small insertion and deletion (indel), and ANNOVAR for variant characterization ^18-20^. Identified variants were filtered through comparison with the disease-associated variants in the known databases (Human Gene Mutation Database (HGMD, 2020.2) and the Online Mendelian Inheritance in Man (OMIM)) and polymorphism databases (dbSNP143, 1000 Genome). Sanger sequencing of the *CRMP1* gene (NM_001014809) was performed to confirm the identified variants in the patients and further family members and performed segregation analysis of the variants with the phenotype.

In family 2, variants were detected by routine WES diagnostics and variant calling using a parent-offspring trio approach as described previously ^21^. Briefly, the exome was captured using the Agilent SureSelectXT Human All Exon v5 library prep kit (Agilent Technologies, Santa Clara, CA, USA). Exome libraries were sequenced on an Illumina HiSeq 4000 instrument (Illumina, San Diego, CA, USA) with 101 bp paired-end reads at a median coverage of 75× at the BGI Europe facilities (BGI, Copenhagen, Denmark). Sequence reads were aligned to the hg19 reference genome using Burrows-Wheeler Alignment (BWA) and variants were subsequently called by the Genome Analysis Toolkit (GATK) unified genotyper, version 3.2-2 and annotated using a custom built diagnostic annotation pipeline. Multi-species sequence alignment were performed using Multialin, PhyloP and PhastCons and the variants disease-causative nature were predicted using the free online tool Mutation Taster (www.mutationtaster.org).

In family 3, trio-based whole-genome sequencing was performed on blood-derived genomic DNA at the Laboratoire de biologie médicale multisites SeqOIA (LBMS SeqOIA, Paris, France). After extraction, DNA samples were sonicated, and libraries were prepared using the NEBNext Ultra-II kit (New England Biolabs). Whole-genome sequencing was performed in paired-end 2×150-bp mode, with a NovaSeq6000 (Illumina). Demultiplexed data were aligned to the genomic reference (hg19). Recalibration and variant calling was performed using GATK4 (Broad Institute). Short variants were annotated with SNPeff (4.3t). Copy Number Variants >4kb were called using CNVnator (v0.4.1) and annotated with AnnotSV (v2.5.1). An average depth-of-coverage of >50x was obtained for both probands, and variants were prioritized according to impact, frequency, and segregation. Sanger sequencing was performed to confirm the identified mutation in the *CRMP1* gene.

### Expression and purification of recombinant *hCRMP1* variants

Subcloned GST-tagged *CRMP1B*-V5 expression plasmids (wildtype, P475L, T313M) were transformed into *E. coli* BL21 (DE3) pLysS cells (BioDynamics Laboratory, Tokyo, Japan). These bacteria were harvested to 200 ml Terrific Broth medium (Invitrogen, Waltham, MA, USA) including 100 mg/ml ampicillin (Sigma, St. Louis, MO, USA). After incubation for 1.5 h at 37 °C, the protein production was induced by 0.1 mM isopropyl β-D-1-thiogalactopyranoside (Sigma) for 18 h at 20 °C. Collected bacteria by centrifugation were suspended in 24 ml homogenization buffer (H buffer; 50 mM Tris/HCl (pH 8.0), 100 mM KCl, 1 mM EDTA, 5% glycerol, 1% NP-40, 1 mM DTT) and then sonicated 4 times for 5 min on ice using Ultrasonic Homogenizer (Microtec Co., LTD., Chiba, Japan). After centrifugation at 12,000 rpm for 30 min at 4 °C, resultant supernatants were mixed with 400 μl of Glutathione Sepharose 4FF resin (Cytiva, Tokyo, Japan) and gently inverted for 24 h at 4 °C. The resins were collected and washed 4 times with 10 ml H buffer, and then washed 5 times with 1 ml cleavage buffer (C buffer; 20 mM Tris/HCl (pH 8.0), 150 mM NaCl, 0.1% NP-40, 1 mM DTT). The pelleted resins were suspended in 100 μl of C buffer including 10 U PreScission protease (Cytiva) and incubated for 24 h at 4 °C with constant agitation. These mixtures were transferred to the filter cups (Vivaclear Mini 0.8 μm PES; Sartorius, Stonehouse, UK), and the flow through fractions were obtained by centrifugation at 10,000 rpm for 1 min at 4 °C. Protein concentrations were determined using the Bradford reagent (Thermofisher scientific, Waltham, MA, USA), and the purified mutants were assessed by SDS-PAGE.

### Determination of dominant-negative effect of the CRMP1 variants on the multimer formation of the wild type

HEK293T cells were co-transfected with pc3.1beta2-V5-*CRMP1B*-wildtype and either one of myc-tagged *CRMP1B* constructs (wildtype, P475L or T313M). After 24h incubation, the cells were harvested by modified C buffer (20 mM Tris/HCl (pH 8.0), 150 mM NaCl, 1% NP-40, 1 mM DTT, protease inhibitor cocktail (cOmplete mini; Roche)), sonicated for 15 min, and centrifuged at 15,000 rpm for 30 min at 4 °C. Electrophoresis of the resultant supernatants under the native and denaturing state were performed as described above. Semi-dry Western transfers were carried out at 15 V for 60 min using the transfer buffer, 50 mM Tris, 40 mM glycine, 20% methanol, 0.1% SDS. In the case of the BN-PAGE, the transferred membranes were incubated in 8% acetic acid for 15 min, washed by ultrapure water, and then dried completely, followed by decolorization using 100% methanol. After blocking by 5% skim milk in TBS-T (20 mM Tris/HCl (pH 7.4), 150 mM NaCl, 0.1% Tween-20) for 60 min, the membranes were reacted with anti-myc (9E10) mAb (1:2,000, FUJIFILM, Cat No. 011-21874) or anti-V5 mAb (1:10,000, Invitrogen (R960−25) RRID: AB_2556564) for overnight at 4 °C. Infrared-fluorescence-(IRDye 680RT; LI-COR, Lincoln, NE, USA) or HRP-conjugated (Dako, Santa Clara, CA, USA) secondary antibodies were utilized for the detection of the first antibodies on the membranes obtained from the native or denatured electrophoresis, respectively.

### Transfection to primary cultured mouse embryonic cortical neurons

Pregnant ICR (RRID:IMSR_TAC:icr) female mice were purchased from Nihon SLC (Shizuoka, Japan). Overdose isoflurane (7 - 8%)/air mixture was inhaled to euthanize the pregnant ICR mice until the loss of breathing. The E15-16 mouse embryos dissected from the pregnant mice were immediately placed in ice-cold PBS and euthanized by decapitation. Cortices dissected from the embryos were treated with 1% trypsin/PBS at 37 °C for 5 min and quenched with the addition of 0.5% trypsin inhibitor. The dissociated neurons were centrifuged at 800 xg for 5 min at 4 °C, washed once, and suspended in DMEM-10% FBS. The cells (1.0 × 10^6^) were transfected with 10 μg of pc3.1beta2-V5 harboring *CRMP1B*-wildtype, -P475L, or -T313M or using an electroporation equipment NEPA21 (NEPA GENE, Chiba, Japan). The condition of electroporation was follows: Poring pulse (275 V, 0.5 ms pulse length, 50 ms pulse interval, 10% decay, +pulse orientation, 2 times); Transfer pulse (20 V, 50 ms pulse length, 50 ms pulse interval, 40% decay, ±pulse orientation, 5 times). The cells were suspended in DMEM-10% FCS and seeded (1 × 10^4^/well) on a 24 well culture-plate coated with 0.05 mg/ml of poly-l-lysine (Wako, Cat No. 163-19091). After overnight incubation, the medium was replaced with Neurobasal medium supplemented with 2% B-27, 2 mM Glutamax, 50 U/ml penicillin, and 50 μg/ml streptomycin (500 μl/well) and incubated 37 °C for 6–7 days.

### Immunocytochemistry of mice cortical cultured neurons

The primary cultured cortical neurons were fixed with 4% paraformaldehyde (PFA)/PBS for 30 min at 25 °C, replaced with PBS, and stored at 4 °C. As the expression of V5-*CRMP1B* constructs was limited, tyramide signal amplification (TSA) system (PerkinElmer, Cat No. NEL700A001KT) was applied for the detection of V5 signal. Briefly, the cells were treated with PBS containing 0.3% H_2_O_2_ for 20 min at 25 °C and blocked with 1:1 mixture of PBS supplemented with 0.1% Triton X-100 (PBST) containing 10% skim milk and Tris-buffered serine supplemented with 0.1% Triton X-100 (TBST) containing 5% normal goat serum (NGS) for 30 min at 25 °C. The cells were washed twice with TBST and incubated with the primary antibody mixture consisting of TBST 2.5% NGS, anti-V5 mouse mAb (1:5000) and anti-MAP2 rabbit pAb (1:5000, Covance (PRB−547C) RRID: AB_2565455) for overnight at 4 °C. The cells were sequentially incubated each for 1h at 25°C with TBST 2.5% NGS containing biotin-conjugated anti-mouse secondary antibody (1:3000, Jackson) and TBST 2.5% NGS containing streptavidin-HRP conjugate (1/500, dilution, PerkinElmer). Then the cells were reacted with tyramide signal amplification mixture for 10min at 25°C and subsequently incubated with TBST 2.5% NGS containing streptavidin-Alexa594 (1:2000) and anti-rabbit secondary antibody-Alexa488 for 2 h at 25 °C. The images of immunostained neurons were captured by an Olympus IX70 microscope equipped with x10 Objective lens and DP74 camera. The neurite outgrowth of V5- or tdTomato-positive neurons was scored with Fiji software (2.0.0-rc-59/1.51n). In each condition, 13 to 20 V5- or tdTomato-positive neurons were analyzed.

## Results

### Phenotype and genotype of index patients

Proband 1 (P1) was born as the second child of non-consanguineous healthy parents of Caucasian descent after an uneventful pregnancy **(Fig 1A, Table 1)**. At delivery a singular umbilical artery was noted. The global development was delayed from infancy on: with sitting at 1.5-2 years-of-age, standing with support at 1-1.5 years-of-age, walking at 2-2.5 years-of-age, first words at 2-2.5 years-of-age. Standardized cognitive tests performed at 5-10 years revealed a moderate intellectual disability with an intelligence quotient (IQ) of 55 at last assessment using the Kaufman Assessment battery for children (K-ABC). She had speech disorder. Behavioral problems included a lack of distance to men and a sexualized behavior. At last assessment at 15-16 years-of-age, the girl had generalized muscular hypotonia with normal reflexes, but fine motor problems with a broad-based gait but no ataxia. When climbing stairs, she showed clear instability. The results of cranial magnetic resonance imaging (MRI) at 1.5 - 4 years as well as that of further work-up (including metabolic tests, electroencephalogram (EEG), ophthalmological assessment, electrocardiogramm, echocardiogramm, abdominal sonography) were normal. Since preliminary genetic tests including chromosome analysis and array-CGH were normal in the index patient (P1), we performed whole exome sequencing (WES) to identify the underlying genetic cause. WES followed by bioinformatic analysis and confirmation with Sanger sequencing revealed the heterozygous *de novo* variant in the *CRMP1* gene c.1766C>T (NM_001014809.2; Chr4 (GRCh37):g.5830253G>A) in the affected child P1 **(Fig 1B)**. At the protein level, this variant leads to an amino acid change at position 589 from proline to leucine: P589L in long isoform, CRMP1A (NP_001014809.1) and P475L in short form, CRMP1B (NP_001304.1) **(Fig 1B)**. The identified variant was not found in 1000Genomes, dbSNP or gnomAD. The CADD score (https://cadd.gs.washington.edu/) was 24.5. The mutation localizes to a highly conserved position, as demonstrated by PhyloP (5.327), PhastCons (1) score and multi-species sequence alignment (**Fig 1C)**. The Provean score of the identified variants predicted to have a deleterious effect (−5.854, cutoff= -2.5).

**Table 1:** Phenotype of patients with *CRMP1* variants.

| Characteristics and Symptoms | Proband 1<br>(Pedigree I) | Proband 2<br>(Pedigree II) | Proband 3<br>(Pedigree III) |
| --- | --- | --- | --- |
| *CRMP1* variant<br>(NM_001014809.2);<br>(NP_001014809.1) | c.1766C>T;<br>p.P589L | c.1280C>T;<br>p.T427M | c.1052T>C;<br>p.F351S |
| Parents | non-consanguineous | non-consanguineous | non-consanguineous |
| Gender | female | male | female |
| Anthropometric data<br>weight (kg)<br>height (cm)<br>OFC (cm) | normal<br>57.1 (0.15 SD)<br>168 (0.43 SD)<br>53.5 (-1.02 SD) | normal<br>N/A | Overgrowth<br>133 (14.2 SD)<br>166.5 ( 2.94 SD)<br>62 (5 SD) |
| Pregnancy, birth, postnatal<br>adaption<br>singular umbilical artery<br>macrosomia<br>fetal fingerpads | normal<br>+<br>-<br>- | normal<br>-<br>+<br>+ | normal<br>-<br>-<br>- |
| Microcephaly (OFC < -2 SD) | - | - | - |
| Macrocephaly (OFC > -2 SD) | - | - | + (5 SD ) |
| Facial dysmorphism | - | - | + |
| Delayed motor development<br>walking unsupported (years) | +<br>2-2.5 | +<br>2-2.5 | +<br>2-2.5 |
| Global muscular hypotonia | mild | mild | mild |
| Deep tendon reflexes | normal | normal | normal |
| Intellectual disability | moderate (IQ 55) | no (IQ 95) | moderate |
| Autism spectrum disorder | - | + | - |
| Behavioral problems<br>lack of distance, sexualized<br>behavior<br>hyperphagia | +<br>+<br>- | +<br>-<br>+ | +<br>-<br>+ |
| Delayed speech and language<br>development<br>(first words spoken) (years) | +<br>(2-2.5) | +<br>(3-3.5) | +<br>(3-3.5) |
| Fine motor problems | + | - | + |
| Other<br>secondary enuresis<br>nocturna<br>pes planus<br>obesity | -<br>-<br>- | +<br>+<br>+ | +<br>-<br>+ |
| EEG results | normal | normal | normal |
| Cranial MRI abnormalities | - | - | - |

+, yes; -, no; IQ, intellectual quotient; N/A, not available; OFC, occipitofrontal circumference; SD, standard deviation.

**Fig 1:**
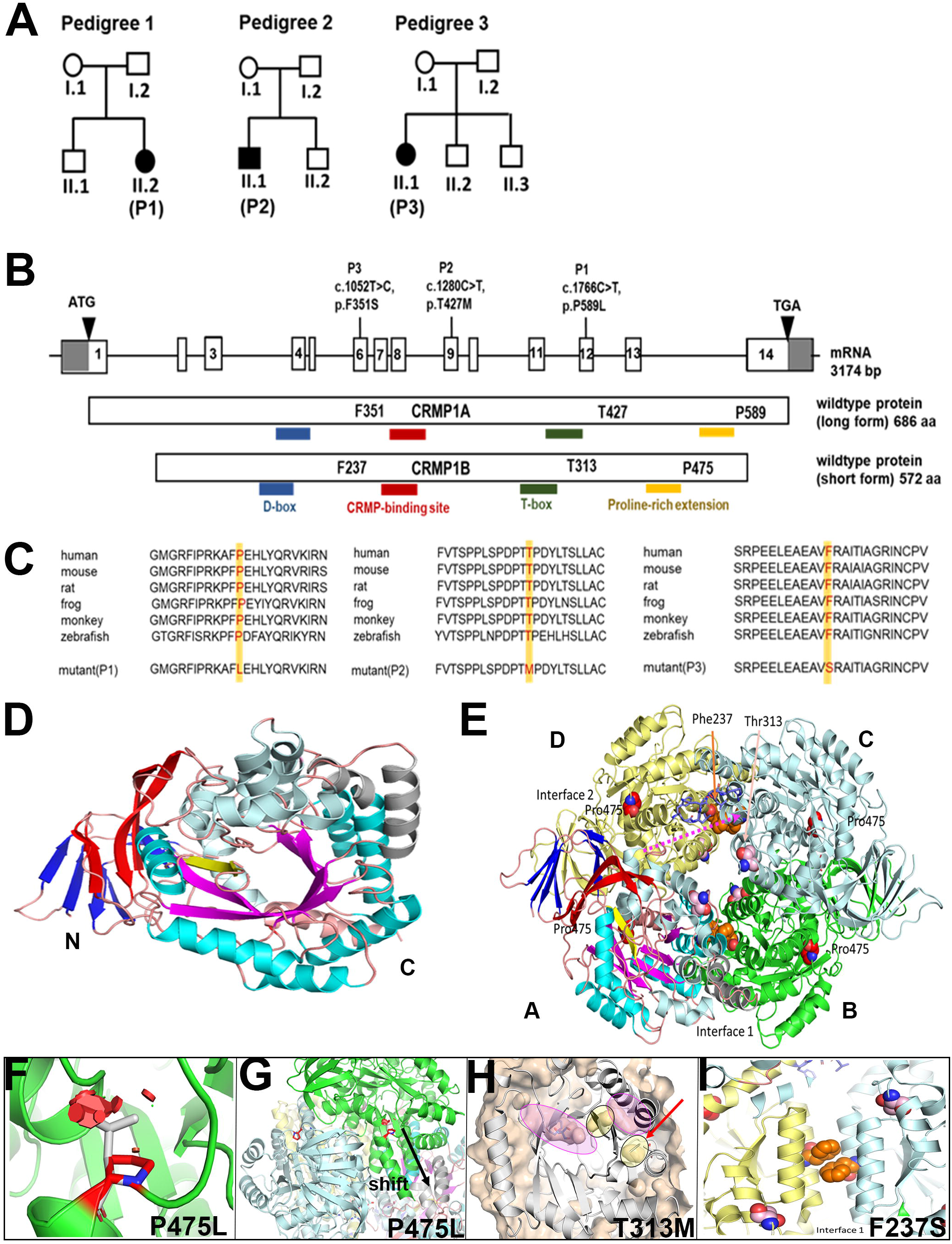
Genotype of patients with variants in *CRMP1*. **A**. Pedigree of index families. **B**. Pictogram representing the CRMP1 cDNA with identified variant of proband 1 (P1) in exon 12 (c.1766C>T, NM_001014809.2) which leads on protein level to an amino acid change of proline to leucine in CRMP1 (CRMP1A-long form (p.P589L, NP_001014809.1) and CRMP1B-short form (p.P475L, NP_001304.1)); the variant in proband 2 (P2) in exon 9 c.1280C>T (NM_001014809.2) leads to an exchange of threonine to methionine (CRMP1A (p.T427M, NP_001014809.1) and CRMP1B (p.T313M, NP_001304.1)); the variant in proband 3 (P3) in exon 6 c.1052T>C (NM_001014809.2) leads to an exchange of phenyl alanine to serine (CRMP1A (p.F351S, NP_001014809.1) and CRMP1B (p.F237S, NP_001304.1)). **C**. Multi-species sequence alignment localizes the variants in the highly conserved area of CRMP1. **D**. The short form CRMP1 monomer is composed of three structural parts, an N-terminally located 7 β-strands forming two β-sheets (depicted in blue), followed by a linker β-strand (yellow) connecting to the central α/β-barrel (cyan/magenta) formed by 7 repeats. Inserted after repeat 4 are 2 additional α-helices (gray) **E**. CRMP1 assembles into tetramers. The relevant sites of mutation T313M, P475L and F237S are indicated as sphere model, with the T313 and F237 located in the central channel in the vicinity of the interaction sites and P475L is locates at the beginning of the C-terminal helix and oriented towards the adjacent molecules. **F**. The mutation P475L reveals serious clashes with neighboring residues (red hexagonals) which may be accounted for by a shift of the helix as shown in (**G). H**. Detailed representation of the structural vicinity of the T313M (yellow) to the ligand-binding cavity (magenta). **I**. Magnified view of interface 1 tilted 90 ° backwards complex with respect to panel E highlighting the arrangement of the two phenylalanines at position 237 from the neighboring units. The exchange of phenylalanine with hydrophobic residues to serine with hydrophilic side chain interferes with the stability of the interaction in interface 1.

Proband 2 (P2) was born as the second child of non-consanguineous parents of Caucasian descent after an uneventful pregnancy and delivery **(Fig 1A, Table 1)**. The boy was macrosomic at birth but had no congenital microcephaly. His motor development was delayed (unsupported walking at 2-2.5 years-of-age) due to congenital mild muscular hypotonia with normal deep tendon reflexes, but no coordination problems. He had bilateral pes planus. A speech delay and language impairment with first words spoken at 3-3.5 years-of-age were diagnosed. He was also diagnosed with an autism spectrum disorder and normal cognitive abilities (IQ 95). At last assessment at 10-11 years-of-age, an obesity associated with hyperphagia was of raising concern, and he had secondary enuresis nocturna. The results of a cranial MRI and EEG were normal. Similarly, through WES followed by bioinformatic analysis, we identified a heterozygous *de novo* variant c.1280C>T in the *CRMP1* gene (NM_001014809.2; Chr4 (GRCh37):g.5841279G>A) in proband (P2) **(Fig 1B)**. This variant leads to an exchange of threonine to methionine at position 427 in the long form, CRMP1A (p.T427M, NP_001014809.1) and at position 313 in the short form, CRMP1B (p.T313M, NP_001304.1) **(Fig 1B)**. The variant affects a highly conserved region of the protein **(Fig 1C)**.

Proband 3 (P3) is the first child of three of a non-consanguineous family of European descent **(Fig 1A, Table 1)**. She was born at gestation week 40 after an uneventful pregnancy, with normal birth parameters (weight: 3.610 kg, height: 50 cm, head circumference: 32.5cm, Apgar score: 10/10). She had developmental delay with not being able to sit alone at 1-1.5 years-of-age and walked without support at 2-2.5 years-of-age. Her speech development was delayed with few words spoken at 2.5-3 years-of-age. She began to gain weight from 1.5-2 years-of-age, and overgrowth was noticed since the 2-2.5 years-of-age. Endocrinological screening, including leptin blood level was normal. There is familial history of obesity on both parental sides, and one of the parent is macrocephalic. She developed severe behavioral issues with temper tantrums, stubbornness, hyperphagia, obsessive-compulsive characteristics and autism spectrum disorder. She was attending medical-educational institute at the age of 8-8.5 years. At the last assessment of 13-14 years-of-age, she had moderate intellectual disability and persistent severe behavioral disorders. Distinctive facial features were low forehead hair insertion, anteverted and large earlobes, broad nasal tip, short philtrum and full lower lips. Genu valgum and hyperlordosis as well as abdominal and dorsal strech marks were noted. Enuresia was noted. Cerebral MRI and abdomino-renal ultrasound were normal. Metabolics and storage disease screening were negative. Array analysis revealed two maternally inherited deletions: a 668 kb deletion at 3q26.31 and a 371kb at 5q23.1, confirmed by genome sequencing and considered as variant of unknown significance. Further analysis through trio-based whole-genome sequencing identified a *de novo* variant in the *CRMP1* gene c.1052T>C (NM_001014809.2; Chr4 (GRCh37):g.5841409A>G) **(Fig 1B)**. The identified variant leads to an exchange of phenyl alanine to serine at position 351 of long form of CRMP1 (CRMP1A (NP_001014809.1)) and at position 237 in short form (CRMP1B (NP_001304.1)) and it is located in the highly conserved region of the protein **(Fig 1C)**. This variant is not found in gnomAD database. *In silico* pathogenicity prediction tools predicts the identified variants to be deleterious (CADD pared: 23.60; REVEL 0.577 (Thresholds > 0.5 Damaging), ClinPred 0.975 (Damaging ≥ 0.5), Mistic: 0.90 (Damaging Threshold ≥ 0.5)). No additional pathogenic variants have been identified by trio genome sequencing, including all known genes involved in neurodevelopmental disorder.

### Effect of identified *CRMP1* variants on protein structure

Since CRMP1 is known to oligomerize to form homotetramers and heterotetramers along with other CRMPs to regulate cellular functions, we determined the effect of the identified human variants on its protein structure using known structures and protein structure prediction tools. The structure of CRMP1 short form, CRMP1B, has been determined at 3.05A resolution^22^ lacking the N-terminal 14 residues and residues 491-572 in their C-terminal region. CRMP1 monomer consists of three structural parts, N-terminal β-strands followed by a linker β-strand connected to the central α/β-barrel **(Fig 1D)**. Several residues contribute to the oligomerization interface of CRMP1 and the quaternary structure of CRMP1 tetramer is shown in **Fig 1E**. The amino acid exchange of P475L, T313M, and F237S are located in the highly conserved region of CRMP1. The P475L is the last residue before the start of the C-terminal helix and the exchange of proline to leucine is predicted to lead to serious clashes with the neighboring residues **(Fig 1F)**. Such spatial constraints may alter the dipole moment of the helix and lead to long-range allosteric effects **(Fig 1G)**. The T313M lies within the region of the α/β-barrel, is oriented towards the inside of the protein and located close to the ligand-binding active site/cavities **(Fig 1H)**. Rearrangements within the protein upon interaction with other CRMPs or molecules could shuffle these cavities for normal functioning. The exchange of threonine to methionine is predicted to prevent rearrangements and to affect the oligomerization as well as other interactions. The F237S is localized directly in the center of interface 1 so the exchange of a heavy hydrophobic sidechain by a hydrophilic short sidechain most likely interferes with the stability of the dimer interaction **(Fig 1I)**. Based on structural simulations, all three variants are predicted to affect the ternary structure of CRMP1 and impact on its oligomerization.

### P475L and T313M mutations affect homo-oligomerization of CRMP1B

To analyze the effect of *CRMP1* variants on its protein levels and cellular function, two variants (CRMP1B-P475L (P1) or -T313M (P2)) were chosen for further functional analysis. We purified the recombinant human CRMP1B-wildtype, -P475L (P1) or -T313M (P2) proteins using the *E. coli* GST-tag expression system. CRMP1B-wildtype showed two major 64 kDa and 60 kDa bands on SDS-denatured gel electrophoresis **(Fig 2A, left lane)**. The 64 kDa and 60 kDa correspond to full-length CRMP1B and to a truncated, C-terminal region cleaved form, respectively. The yield of purified T313M (TM) or P475L (PL) was less than that of wildtype in the same condition **(Fig 2A, middle and right lanes**). This finding may be due to lower expression and/or aggregation of the mutant proteins in *E. coli*. As T313M and P475L mutated residues are positioned close to the dimer/tetramer interface of CRMP1B, these mutations may affect homo-oligomerization of CRMP1. We therefore examined the oliogomerization of recombinant CRMP1 preparation with Blue-Native gel electrophoresis **(Fig 2B)**. A broad band of CRMP1B-wildtype was present at high (720 kDa) but residual amount of monomeric form was detected at low molecular weight (70 kDa) on Blue-Native gel. The 720 kDa band probably represents the homo-oligomerization of CRMP1B-wildtype under the native condition. In contrast, CRMP1B-T313M and -P475L showed the increased monomeric 70 kDa band, suggesting weakened self-association of these mutants.

**Fig 2.**
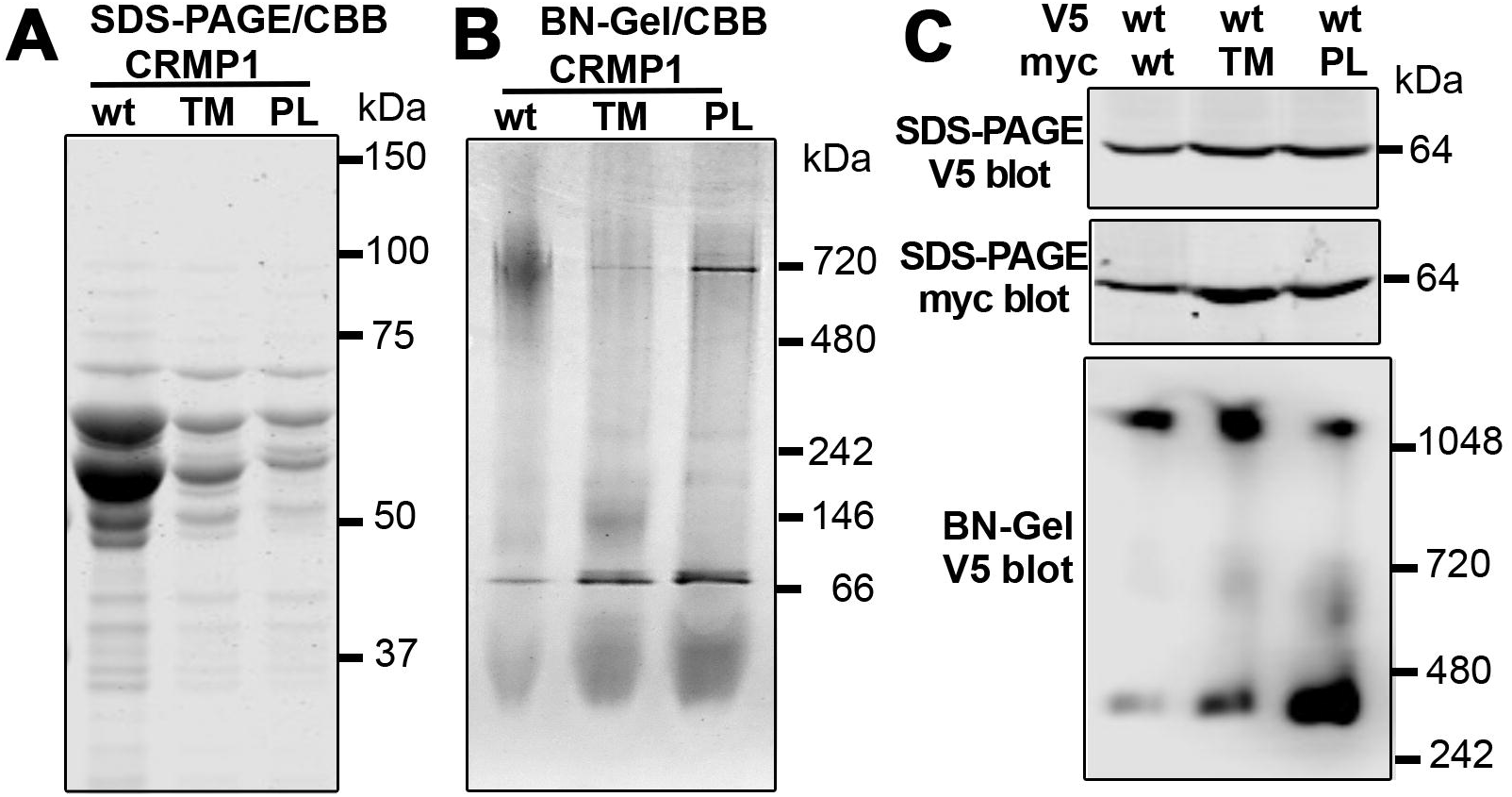
Attenuated oligomer formation of CRMP1-P475L and CRMP1B-T313M mutants. **A**. Purified CRMP1B-wildtype, -T313M, and -P475L recombinant proteins on SDS-PAGE stained with CBB. GST-tagged CRMP1 were expressed in *E. coli* and purified through the binding to glutathione resin and the digestion with PreScission protease. Equal volume (2.5 μl) of the purified specimens, which were prepared in the same condition, were loaded on the gel. The yield of the mutants was less than that of CRMP1-wildtype as shown in panel. Two major 64 kDa and 60 kDa bands in the preparations are full-length and truncated form, respectively. **B**. Estimation of the multimeric state under native condition. Bacterially expressed and purified CRMP1B specimens (5 g) were electrophoresed on NativePAGE Novex 4-16% Bis-Tris gel in the presence of CBB-G250. The gel was stained with CBB-R250 to visualize separated proteins. CRMP1B-wildtype exhibited major 720 kDa and minor 70 kDa bands, which respectively represent homo-oligomer and monomer. In contrast, CRMP1B-T313M and -P475L showed predominant 70 kDa bands, suggesting that the mutations may interfere with the homo-oligomerization of CRMP1. **C**. Dominant negative effect of T313M and P475L mutant on the oligomerization of CRMP1B-wildtype. HEK293T cells co-expressing V5-CRMP1B-wildtype and either one of myc-CRMP1B-wildtype, - T313M, or -P475L were analyzed. Anti-V5 and anti-myc immunoblot analyses of the specimens subjected to SDS-PAGE showed that the expression level of V5-CRMP1 and myc-CRMP1 in each condition was similar extent (upper and middle panels). Anti-V5 immunoblot analysis of these specimens on Blue-Native gel electrophoresis is shown (lower panel). Co-expression of V5-CRMP1B-wildtype and myc-CRMP1B-wildtype exhibited major 1100 kDa and minor 360 kDa V5-reactive bands. In contrast, co-expression of myc-CRMP1B-T313M or -P475L increased the lower (360 kDa) V5-reactive band. This suggests that co-expression of these mutants may interfere with multimer formation of V5-CRMP1B-wildtype. Abbreviations: CRMP1B-wildtype, wt; CRMP1B-T313M, TM; CRMP1B-P475L, PL

As these mutations were found in the heterozygous state in the affected patients, CRMP1B-T313M and/or -P475L expression might perturb the oligomerization of CRMP1B-wildtype in a dominant-negative manner. We therefore next co-expressed V5-*CRMP1B*-wildtype together with either myc-*CRMP1B*-wildtype, -T313M, or -P475L in HEK293T cells. As shown in **Fig 2C** upper and middle panels, co-expression of myc-*CRMP1B*-T313M or -P475L did not alter the expression level of V5-CRMP1B-wildtype on SDS-denatured gel electrophoresis. Anti-V5 immunoblot analysis of these specimens on Blue-Native gel electrophoresis (**Fig 2C**, lower panel) revealed that the co-expression of myc-*CRMP1B*-wildtype showed major 1100 kDa and minor 360 kDa bands of V5-CRMP1-wildtype. In contrast, co-expression of myc-*CRMP1B*-T313M or -P475L increased the lower (360 kDa) V5-CRMP1B-wildtype, suggesting that these mutants potentially impede multimer formation of CRMP1B-wildtype. As V5-immunoreactive signal at higher molecular size (1100 kDa) in P475L was remarkably decreased than that in T313M, P475L may exhibit stronger dominant negative effect.

### P475L and T313M mutations attenuate neurite outgrowth of cortical neurons

We next asked whether the ectopic expression of *CRMP1B* mutants in primary cultured murine cortical neurons affect the neuronal development. Dissociated E15 mouse cortical neurons were electroporated with the expression vector harboring either V5-*CRMP1B*-wildtype (wt), -T313M (TM), -P475L (PL), or tdTomato. The cells were seeded onto PLL-coated culture dishes and grown for 6-7 days. After fixation, the cells were immunostained with anti-V5 and anti-MAP2 antibodies. We found that the longest primary neurites of V5-*CRMP1B*-T313M or -P475L transfected neurons were shorter than those of V5-*CRMP1B*-wildtype (wt) or tdTomato transfected cells **(Fig 3A-D)**. We then measured the length of the longest primary neurite from V5-or tdTomato-positive neurons in each condition. The longest primary neurites expressing *CRMP1B*-T313M or -P475L were 40-50% shorter than those expressing CRMP1B-wildtype (wt) **(Fig 3E)**. As the longest primary neurites of the cultured cortical neurons is thought to be axons, these CRMP1B mutants may interfere with the oligomerization of CRMP1 in turn to attenuate the outgrowth of the cortical axons.

**Fig 3.**
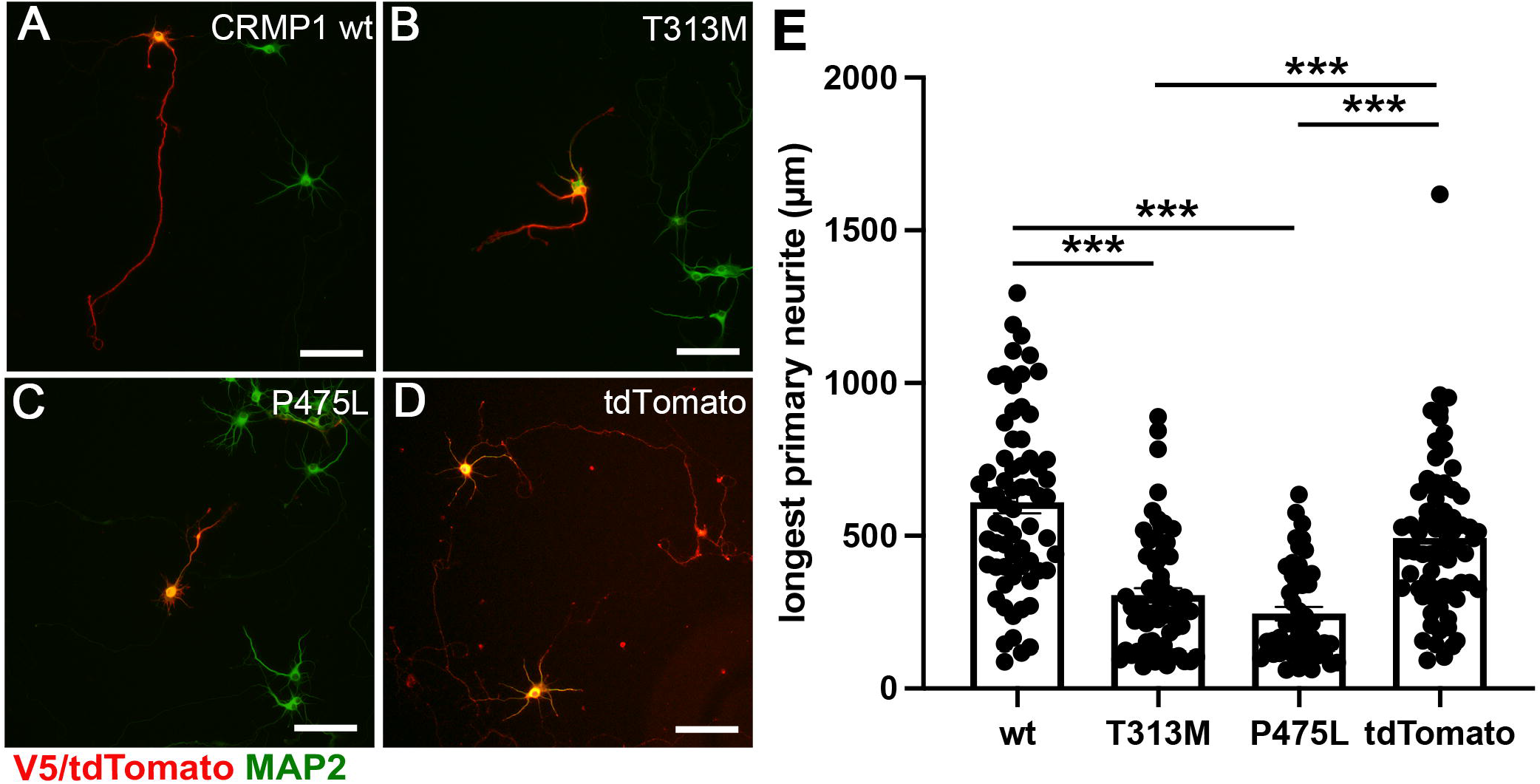
Attenuated neurite outgrowth by the ectopic expression of CRMP1B-P475L and CRMP1B-T313M mutants. **A-D**. Representative images of the neurons expressing V5-CRMP1B-wildtype (A), -T313M (B), -P475L (C), or tdTomato (D). Transfected neurons were visualized by anti-V5 immunostaining or tdTomato expression (red) and anti-MAP2 immunostaining (green). The longest primary neurites of the neurons expressing V5-CRMP1B-T313M or -P475L were shorter than those of the neurons transfected with V5-CRMP1B-wildtype or tdTomato. Scale bars, 100 μm. **E**. Longest primary neurite length. The length of the longest primary neurite from V5- or tdTomato-positive neurons was scored in each condition. The graph represents average ± SEM with individual values from four independent experiments. The number of examined neurons in each condition is CRMP1B-wildtype, 66; -T313M, 64; -P475L, 49; tdTomato, 73. Data were analyzed by one-way ANOVA followed by Tukey’s post-hoc test. \*\**p*< .01, \*\*\**p* <0.001.

## Discussion

In this study, we report the human phenotype associated with heterozygous *CRMP1* variants in three affected children of unrelated non-consanguineous pedigrees. All the patients have a neurodevelopmental disorder with motor delay and muscular hypotonia. While patient P1 has moderate intellectual disability and behavioral abnormalities, P2 was diagnosed with an autism spectrum disorder but a normal cognitive profile and P3 presented with moderate intellectual disability and autism spectrum disorder.

We showed that those human *CRMP1* variants have a dominant-negative effect and impact the oligomerization of CRMP1 proteins. CRMP1 exists as homo- or hetero-tetramers to interact with various signaling molecules and to link microtubules to subcellular structures and regulate cytoskeletal dynamics.^5,23^ In line with this, CRMP1 colocalizes to the mitotic spindles and centrosomes, indicating of playing a key role in mitosis and cell cycle progression.^24^ Defective cell cycle, abnormal mitotic spindle and centrosomes are the common key pathomechanism underlying several neurodevelopmental disorders.^25^

Dysregulation of other CRMPs that have been linked to human disease has been shown to affect specific cellular processes associated with the clinical phenotype. For example, mutant CRMP5 impedes the ternary complex formation with microtubule-associated protein 2 (MAP2) and beta-III-tubulin, thereby causing a defective inhibitory regulation on neurite outgrowth and dendritic development and thereby contribute to the brain malformation phenotype.^13^ MAP2 has been already reported to play a key role in neurite outgrowth and synaptic plasticity through its role in microtubule stabilization and cytoskeletal association.^26,27^ Also, mutant CRMP4 was shown to affect axonal growth and survival of motor neurons and thereby contribute to the amyotrophic lateral sclerosis phenotype.^14^ In this study we showed that the identified *CRMP1* variants affect neurite outgrowth, which is a characteristic phenotype associated to many neurodevelopmental/psychiatric disorders.^28^ Our finding is in line with the reduced neurite outgrowth phenotype and impaired long-term potentiation in the *Crmp1*^*-/-*^ mice.^9^ Mechanistically, these cellular processes are regulated in a spatiotemporal manner through CRMP1 phosphorylation by several kinases such as Cdk5, Rho/ROCK or GSK3, and any disturbances in these kinases or mutation of specific residues culminates in the abnormal brain development.^6,8-11,29^ In the case of *Cdk5*^-/-^ mice, abnormal dendritic spine morphology was shown in cortical neurons as it was in *Crmp1*^-/-^ mice. Intriguingly, both mouse models show schizophrenia-like behavior, indicating the commonly shared cellular mechanism for dendritic development.^9,30^ In the context of neuronal development, defective phosphorylation of tyrosine residues in CRMP1 by Fyn-mediated Reelin signaling impairs cortical neuron migration.^6^ Over the last two decades, new phosphorylation specific sites in CRMP1 are being reported and recently, phosphorylation of Tyr504 residue by Fyn has been shown to play an important step in Sema3A-regulated dendritic development of cortical neurons.^31^ As a future perspective, it will be interesting to find other *CRMP1* variants and elucidate the role of specific residues in various neuronal process and associate with the phenotypic spectrum of neurodevelopmental disorder.

In conclusion, we report for the first time human *CRMP1* mutations, link them to a human neurodevelopmental disease and highlight underlying pathomechanisms. Our report adds *CRMP1* to list of other *CRMP* genes linked to neurological disorders and underlines the important role of CRMP1 in the nervous system development and function.

## Data availability

All methods applied in this project are described in detail in the ‘Materials and Methods’section and the original data are presented as source file

## Acknowledgements

The authors thank the index families for their participation in this study. We acknowledge Petra Bittigau, Akosua Sarpong for attending the patients; Anna Tietze for the analysis of cranial MR images; Sabrina Pommer and Jessica Fassbender for technical assistance; Piotr Neumann for assistance in data interpretation and lively discussions; Dr. Tanabe Kenji and Dr. Fumitoshi Saitoh for electroporation equipment. We acknowledge the funding resources for this study: German Research Foundation (SFB665, SFB1315, FOR3004, AMK), the Sonnenfeld Stiftung (AMK, KLM), the Berlin Institute of Health (BIH, CRG1, AMK), the Charité (AMK, ER, LLB, KLM), and by grants-in-aid for Scientific Research from the Japan Society for the Promotion of Science (JSPS) (16K07062) (FN).

## Author contribution

AMK and FN were responsible for project conception. AMK, MV, BD analyzed and interpreted patient’s clinical data. LLB and KLM contributed clinical information by gathering clinical information and written informed consents. HH, NL, APAS, JL, TR performed WES and bioinformatics data analysis. ER performed Sanger sequencing and segregation analysis. NA, KT and FN performed functional experiments. AD performed structural prediction analysis. ER and AMK drafted the manuscript that was revised and accepted by all coauthors.

